# Comparing Pathway-Informed Polygenic Risk Score Strategies: A multi-cohort evaluation of Amyloid-β

**DOI:** 10.64898/2026.05.25.26354071

**Authors:** Xiyuan Zhang, Benjamin Goudey, Simon M. Laws, Colin L. Masters, Timothy Baldwin, Noel G. Faux

## Abstract

**Objective:** To systematically evaluate pathway-informed polygenic risk score (PRS) strategies and determine which approaches most effectively leverage biological annotations for risk prediction, using brain amyloid-β (Aβ) positivity as a case study.

**Methods:** We systematically benchmarked approaches for integrating pathway information into PRSs construction to predict brain Aβ positivity. Using two cohorts, the Alzheimer’s Disease Neuroimaging Initiative (ADNI, n = 969) and Australian Imaging, Biomarkers and Lifestyle (AIBL, n = 251), we compared Apolipoprotein E (*APOE*) genetic risk score (GRS), clumping and thresholding (C+T) PRS, pathway-guided single nucleotide polymorphism (SNP) selection PRS, and pathway-specific PRSs ensembled via machine learning. Pathways were derived from manually curated literature or from pathway databases via Functional Mapping and Annotation (FUMA).

**Results:** In cross-validation on the ADNI cohort, pathway-informed PRS using a narrow-set of pathways to guide SNP selection (PathPRS-SNP_Lit_ without *APOE* locus) significantly outperformed the standard PRS model (median AUC = 0.742, p = 0.006) and the APOE locus model (median AUC = 0.736, p = 5.1 × 10^-5^) based on the Mann–Whitney U test, achieving a median AUC of 0.763. This model showed enhanced ability to identify subgroups within the 10% lowest-and highest risk groups compared to the current standard of *APOE* locus alone (odds ratio = 0.67, 95% CI: 0.56–0.81; and OR = 13.23, 95% CI: 10.23–17.11), highlighting its clinical potential. Using a focused set **l**iterature-curated pathways outperformed using a broader set of database-derived pathways across configurations. When contrasting strategies for aggregating information across pathways, we observed that using pathways to guide selection of SNPs and then building a single PRS performed comparably to building PRS for each pathway and using machine learning (ML) to aggregate these, though the latter enabled pathway-level interpretability. **S**imilar trends were observed in the external AIBL validation dataset

**Interpretation:** Pathway-informed PRS can meaningfully improve genetic risk enrichment for Aβ positivity beyond APOE and standard C+T approaches, provided pathway definitions are carefully curated. The choice of pathway source has the strongest impact on predictive performance with aggregation strategies or ML model choice having far less impact. Our findings highlight the utility of literature-curated, pathway-informed PRSs for Aβ prediction and offer practical guidance for pathway-informed PRS construction in other polygenic traits.

## INTRODUCTION

Alzheimer’s disease (AD) is a progressive neurodegenerative disease characterized by the presence of amyloid beta (Aβ) aggregates, also known as Aβ plaques, in the brain. It is now established that Aβ plaque occurs decades prior to presenting clinical symptoms like severe cognitive decline.^1^ The Apolipoprotein E (*APOE*) gene is a key determinant of AD risk with three alleles: □2, □3, and □4. *APOE* □4 significantly increases AD risk in a gene dose-dependent manner, elevating the risk by up to 15-fold in homozygotes.^2^ While *APOE* represents the strongest genetic risk factor for late-onset AD, numerous genome-wide association studies (GWAS) have identified many additional loci that contribute to disease susceptibility, including *CLU*, *BIN1*, *PICALM*, and *CR1*, highlighting the highly polygenic architecture of the disease.^3^

To capture the polygenic nature of AD, several approaches have been proposed to integrate genetic variants beyond *APOE*. While *APOE* confers substantial genetic risk, it is not the only locus associated with AD. Numerous GWAS and meta-analyses have identified additional single nucleotide polymorphisms (SNPs) associated with AD risk, enabling the construction of polygenic risk scores (PRS). One widely-used approach is to combine the effect sizes of SNPs identified as associated with a disease in GWAS. The clumping and thresholding (C+T) method is a widely used approach for constructing polygenic risk score (PRS). C+T clumps variants based on linkage disequilibrium (LD), selects SNPs by a GWAS *p* threshold, and aggregates their effect sizes to compute the PRS.^4^ Numerous studies have explored the use of C+T to improve the prediction of AD-related phenotypes.^5–8^ However, it remains uncertain whether C+T can enhance the prediction of abnormal Aβ deposition beyond the influence of *APOE* alone.^5,8,9^

A key limitation of standard C+T PRS is its limited incorporation of biological knowledge. It selects SNPs solely based on p-value thresholds and LD pruning, without leveraging pathway-level information.^10,11^ Recent studies have begun to address this gap by integrating pathway-specific biological annotations into PRS construction, with mixed findings. Darst et al.^8^ reported that selected pathway-informed PRSs were significantly associated with Aβ deposition and, in some cases, outperformed genome-wide PRS. In contrast, Leonenko et al.^5^ found that pathway-level PRSs offered no additional predictive value beyond genome-wide PRS for Aβ positivity. Thus, it remains unclear how different strategies for incorporating pathway information into PRS influence predictive performance and generalizability, particularly when evaluated in specific applications such as Aβ prediction. Moreover, both studies evaluated each pathway independently rather than considering their joint effects. Given that AD involves multiple interacting molecular pathways,^12^ such separate analyses may fail to fully account for disease risk.

Building on these insights, recent research has proposed using machine learning to combine multiple pathway-specific PRS into a single integrative model.^13^ This study has relied on a single machine-learning approach rather than systematically evaluating a broader range of models, limiting understanding of how different algorithmic choices influence performance. While such methods have been shown to outperform standard PRS in certain complex traits, their utility in Aβ prediction remains uncertain, particularly when comparing different sources of pathway definitions (e.g., databases vs. literature).

In this study, we present a systematic evaluation of pathway-informed PRS (PathPRS) strategies, using Aβ positivity as a representative use case. We specifically compared literature-curated pathways (PathPRS_Lit_) to database-derived pathways (PathPRS_DB_). Each generates multiple pathway-specific PRSs, which were then aggregated into a score using a range of machine-learning models. We benchmarked these approaches against conventional C+T PRS and the *APOE* genetic risk score (GRS), using data from two cohorts: the Alzheimer’s Disease Neuroimaging Initiative (ADNI) for in-domain validation using nested cross-validation, and the Australian Imaging, Biomarkers and Lifestyle (AIBL) for external validation.

## METHODS

### Datasets

This study analysed two longitudinal studies of AD: the Alzheimer’s Disease Neuroimaging Initiative (ADNI) ^15^ and Australian Imaging, Biomarkers and Lifestyle (AIBL)^14^ studies. We constrained the analysis to only consider those individuals for whom both genomic measurements from SNP arrays and brain Aβ deposition from positron emission tomography (PET) imaging had been collected.

Both studies included individuals who were cognitively unimpaired, had mild cognitive impairement or had a clinical diagnosis of Alzheimer’s disease dementia. Baseline characteristics of these studies are described further in Table 1.

**Table 1:**
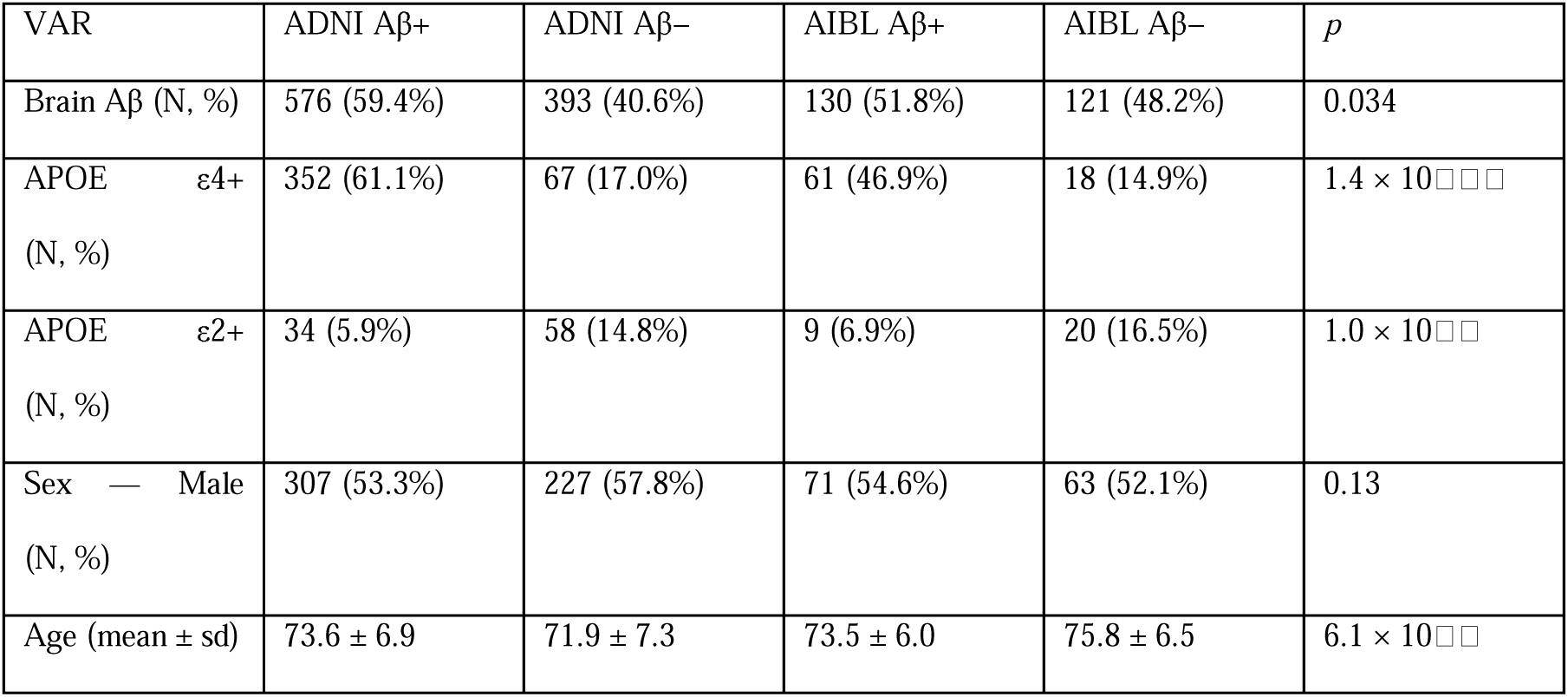
Demographics of ADNI and AIBL datasets, stratified by Aβ status. Values are counts and proportions (in brackets), except for age which shows mean and standard deviations. The p-value indicates whether significant differences were observed across datasets using two-way ANOVA for age and chi-squared test otherwise. Aβ: Amyloid beta.

### Genotyping and Quality Control

In ADNI, brain Aβ deposition was measured via PET with the AV45 (florbetapir) tracer, adhering to standardized acquisition protocols as detailed in the ADNI methods documentation (http://adni.loni.usc.edu/methods/documents/). Brain Aβ deposition was expressed as the mean standardized uptake value ratio (SUVR) across the frontal cortex, anterior cingulate, precuneus, and parietal cortex regions, normalized to the cerebellar reference region.^16^ A standardized centiloid scale was employed to harmonize brain Aβ measurements across datasets.^17^ Brain Aβ positivity was defined as a centiloid greater than 20.^17^ In AIBL, brain Aβ deposition was similarly assessed via PET imaging using multiple tracers, including AV45, Pittsburgh Compound B, NAV4694, flutemetamol, and florbetaben.^18^ Aβ positivity in AIBL was defined by a centiloid value greater than 20 to align with ADNI measurement.

### Genotyping and Quality Control

Individuals in ADNI were genotyped with the Illumina Omni 2.5M BeadChip,^19^ while all individuals in AIBL were genotyped using the Axiom Precision Medicine Diversity Array.^20^ To harmonize SNPs across the two datasets, both datasets were imputed using the Michigan Imputation Server^21^ (https://imputationserver.sph.umich.edu) with Minimac4 as the imputation software, Eagle2^22^ for phasing, and 1000 Genomes dataset^23^ as the reference panel with all results aligned to the GRCh37/hg19 assembly.

Quality control (QC) of the genetic data was conducted using PLINK v1.9.^24^ Following standard quality control recommendations,^4^ SNPs with a missingness rate greater than 0.02, a Hardy-Weinberg equilibrium *p* less than 1×10^−6^, or a minor allele frequency below 0.01 were excluded from further analysis. Prior to QC, the ADNI dataset comprised 1,549 individuals and 26,056,934 SNPs, while the AIBL dataset included 276 individuals and 41,324,331 SNPs. Following QC, 1,549 individuals and 8,091,788 SNPs were retained in the ADNI dataset, and 276 individuals and 8,325,288 SNPs were retained in the AIBL dataset for downstream analysis. A total of 6,676,990 SNPs overlapped between the two datasets.

### PathPRS: A flexible framework for constructing pathway informed PRS

**Fig 1:**
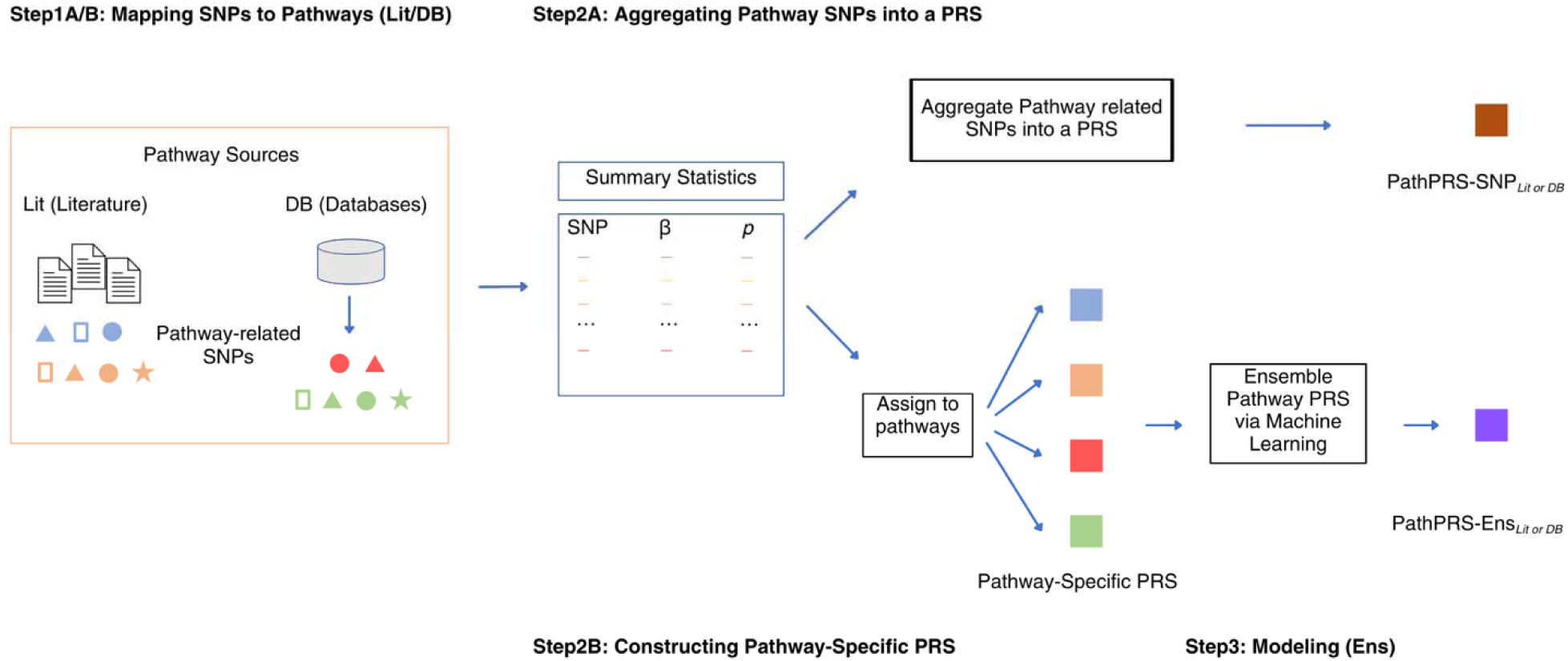
Pipeline for constructing PathPRS. Steps labeled A and B correspond to the two alternative branches of Pat PRS construction: PathPRS-SNP (A) and PathPRS-Ens (B). PathPRS-SNP Branch (2 Steps): Step 1A: SNPs are mapped to pathways using literature-curated (Lit) or database-derived (DB) sources. Step 2A: Pathway-related SNPs are aggregated into a single PRS score, resulting in PathPRS-SNP_Lit/DB_. PathPRS-Ens Branch (3 Steps): Step 1B: SNPs are mapped to pathways. Step 2B: SNPs are grouped by pathway, and pathway-specific PRS are constructed. Step 3B: These pathway-level PRS are input into a machine learning model to generate the final score PathPRS-Ens_Lit/DB_.

### Mapping SNPs to Pathways

Pathways were collected using two primary approaches: a literature-based approach and a database-based approach (Fig 1).

For the literature-based approach, we performed a literature search in major academic databases (i.e., PubMed and Web of Science) using keywords Alzheimer’s disease, pathway, and PRS to find studies that leveraged pathway--informed PRS to predict AD. From this search, we identified five key papers^5,8,25–27^ that not only investigated individual pathway specific PRS but also publicly disclosed “pathway-SNP” mappings (e.g., in supplemental materials), (Supplementary Table S1). Together, these studies provided a total of 32 pathways and 3,003 SNPs for further analysis.

As a complementary approach, we used Functional Mapping and Annotation (FUMA),^28^ an online platform designed for the functional annotation and prioritization of genetic variants from GWAS summary statistics. Based on SNP2GENE and GENE2FUNC analyses in FUMA using International Genomics of Alzheimer’s Project (IGAP) summary statistics,^29^ we identified 275 pathways and 3,120 SNPs that were significantly enriched (FDR < 0.05).

### Pathway-Specific PRS Construction

Both the literature and pathway database approaches result in a set of SNPs for each pathway. As illustrated in step 2 of Fig 1, these pathway-related SNP sets were then mapped to IGAP summary statistics, providing effect sizes and *p* for their relationship with AD.

Based on this, the PRS for an individual pathway *p* is defined as:

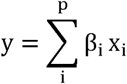

where β*_i_*is the effect size for SNP *i* recorded in the IGAP summary statistics, and x_i_ is the dosage of SNP *i*; *APOE* features were also included (*APOE* □2 genotype, *APOE* □4 genotype) in each PRS.

### Machine Learning Models and Training

To effectively integrate the pathway-specific PRS, we applied a machine learning model designed to generate an aggregated prediction score. In this process, we took each individual pathway-specific PRS as an input feature to the machine learning model, allowing the model to capture potential interactions between pathways.

Multiple types of models were considered to perform this final aggregation, namely: Linear Regression, Lasso, Elastic Net, support vector machine (SVM) with a linear kernel, SVM with a Radial Basis Function (RBF) kernel, Random Forest, and a Multi-Layered Perceptron (MLP). A grid search with nested cross-validation was employed to optimize model hyperparameters (Supplementary Table S2). The MLP model was trained using the Adam optimizer,^30^ with a dropout rate of 0.5 applied to each layer, and LeakyReLU^31^ as the activation function for all layers, except the final layer, where a sigmoid activation function was employed for binary classification. All models except for the MLP were implemented using scikit-learn version 1.5.1;^32^ the MLP was implemented using PyTorch version 2.4.0.^33^ Mean squared error loss was used to train all models to ensure a consistent optimization objective across models, enabling fair comparison of their predictive performance.

#### PathPRS-SNP and PathPRS-Ens

For PathPRS, we developed two complementary models: PathPRS-SNP and PathPRS-Ens.

The PathPRS-Ens framework takes pathway-specific PRSs as inputs, enabling the model to better capture interactions across pathways and to quantify their relative contributions to prediction performance. Depending on the source of pathway definitions, this model is referred to as PathPRS-Ens_Lit_ (literature-curated pathways) or PathPRS-Ens_DB_ (database-derived pathways).

To specifically assess the impact of pathway-informed SNP selection, we constructed a PathPRS-SNP framework that uses the union of all SNPs mapped to AD-related pathways. For each individual, we computed a single PRS by summing effect-size--weighted allele dosages of all pathway-related SNPs, without pathway--level grouping. This resulting score served as a single input feature to the model. Depending on the pathway source, this is denoted as PathPRS-SNP_Lit_ or PathPRS-SNP_DB_.

Within the PathPRS framework, the dosages of *APOE* □ 2 and □ 4 genotypes were included as covariates.

### Baseline Methods

We constructed two baseline methods to understand the performance improvement of the PathPRS approach:

1. *APOE*: This model was a linear combination of the *APOE* □ 2 genotype and *APOE* □ 4 genotype dosages for each individual. The linear combination of two ternary-valued features (for 0, 1, or 2 copies of each genotype) was determined by an Elastic Net model.
2. C+T: A standard C+T model was constructed based on summary statistics from IGAP. SNPs were pruned for LD with an r^2^ threshold of 0.1 and a window size of 1,000kb. The *p* threshold was treated as a hyperparameter in nested cross-validation, selected from 13 options ranging from 0.5 and 0.1 to 1 × 10^−12^.

### Performance Evaluation and Statistical Analysis

The models were trained and evaluated on the ADNI dataset, using 10×10 nested cross-validation, with 10 repeats of stratified cross-validation. Statistical significance in the Area Under the Receiver Operating Characteristic curve distributions across models was determined using a Mann–Whitney U test. All *p* values were adjusted for multiple comparisons using the Benjamini–Hochberg procedure to control the False Discovery Rate. When conducting external validation in AIBL, AUCs of the models were compared using the DeLong test.^34^ To further quantify decile-wise effect differences between models, we performed a fixed-effects meta-analysis using the DerSimonian-Laird method^35^ with continuity correction, assessing both risk differences and odds ratios. In all analyses, a significance threshold of 0.05 was used and p-values were corrected for multiple comparisons using the False Discovery Rate.

### Code availability

Python codes is provided in GitHub (https://github.com/EmmaUoM/pathway_prs) for the PathPRS model training and baseline implementations.

## RESULTS

### Data characteristics

We observed significant differences between the ADNI and AIBL cohorts in several key demographic and genetic variables (Table 1). Notably, the proportion of Aβ-positive individuals differs significantly between the two cohorts, indicating a potential difference in disease burden or recruitment strategies.

Genetic differences were also evident. The prevalence of *APOE* [4 carriers among Aβ-positive individuals was significantly higher in ADNI than in AIBL. Similar trends were observed for *APOE* [2 carriers and across Aβ-negative subgroups. These differences in genetic risk factor distributions suggest different cohort compositions that may affect model generalizability.

In terms of age, Aβ-negative individuals in AIBL were significantly older on average than their counterparts in ADNI, further contributing to demographic heterogeneity.

These inter-cohort differences, particularly in *APOE* genotype frequencies and age distributions, may influence model performance and limit generalizability when applying models trained on ADNI to AIBL. Accordingly, it is important to interpret cross-cohort validation results considering these baseline differences.

### PathPRS Enhances A**β** Prediction Beyond All PRS Baselines

Among the baseline methods, PathPRS-SNP_Lit_ with *APOE* locus excluded achieved the highest median AUC (0.763) (Fig 2). When modeled using Elastic Net, PathPRS-ENS_Lit_ remained significantly better than *APOE* (median AUC: 0.758 vs. 0.736; Mann–Whitney U test, *p* = 0.002), consistent with the results observed from the MLP model. Notably, PathPRS-SNP_Lit_ with *APOE* locus excluded significantly outperformed the conventional C+T PRS (median AUC: 0.763 vs. 0.742, *p* = 0.006). In contrast, pathways derived from databases, PathPRS-SNP_DB_ and PathPRS-ENS_DB_, showed lower performance (median AUC: 0.742 and 0.709, respectively). These results suggest that literature-based, pathway-informed SNP selection enhances brain Aβ prediction beyond what is achieved by the widely-used C+T approach. Furthermore, the comparable performance between PathPRS-SNP_Lit_ and PathPRS-ENS_Lit_ (median AUC: 0.759 vs. 0.758) indicates that this improvement holds both when pathways were modeled individually and when SNPs across all pathways were aggregated (Supplementary Fig S1).

**Fig 2:**
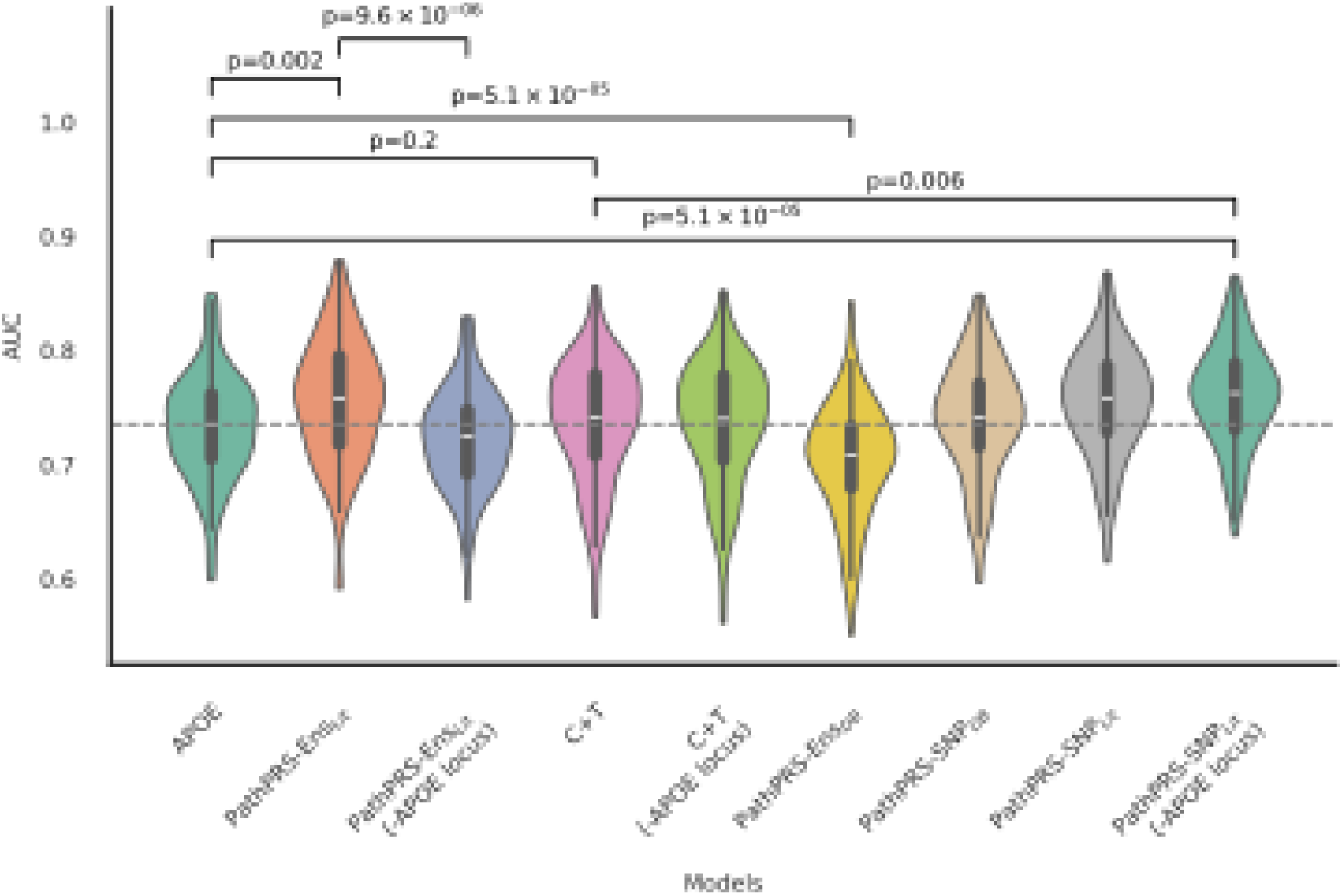
AUC distributions from 10 repeats of 10-fold stratified cross-validation are shown. Elastic Net models were trained using different PRS inputs on the ADNI dataset. Significance of difference in AUC distributions was compared using the Mann-Whitney U test after Benjamini-Hochberg correction. Gray dashed lines represent the median AUC of the *APOE* model. Numbers below each violin indicate the median AUC for that model. Area Under the Receiver Operator Curve = AUC.

**Fig 3:**
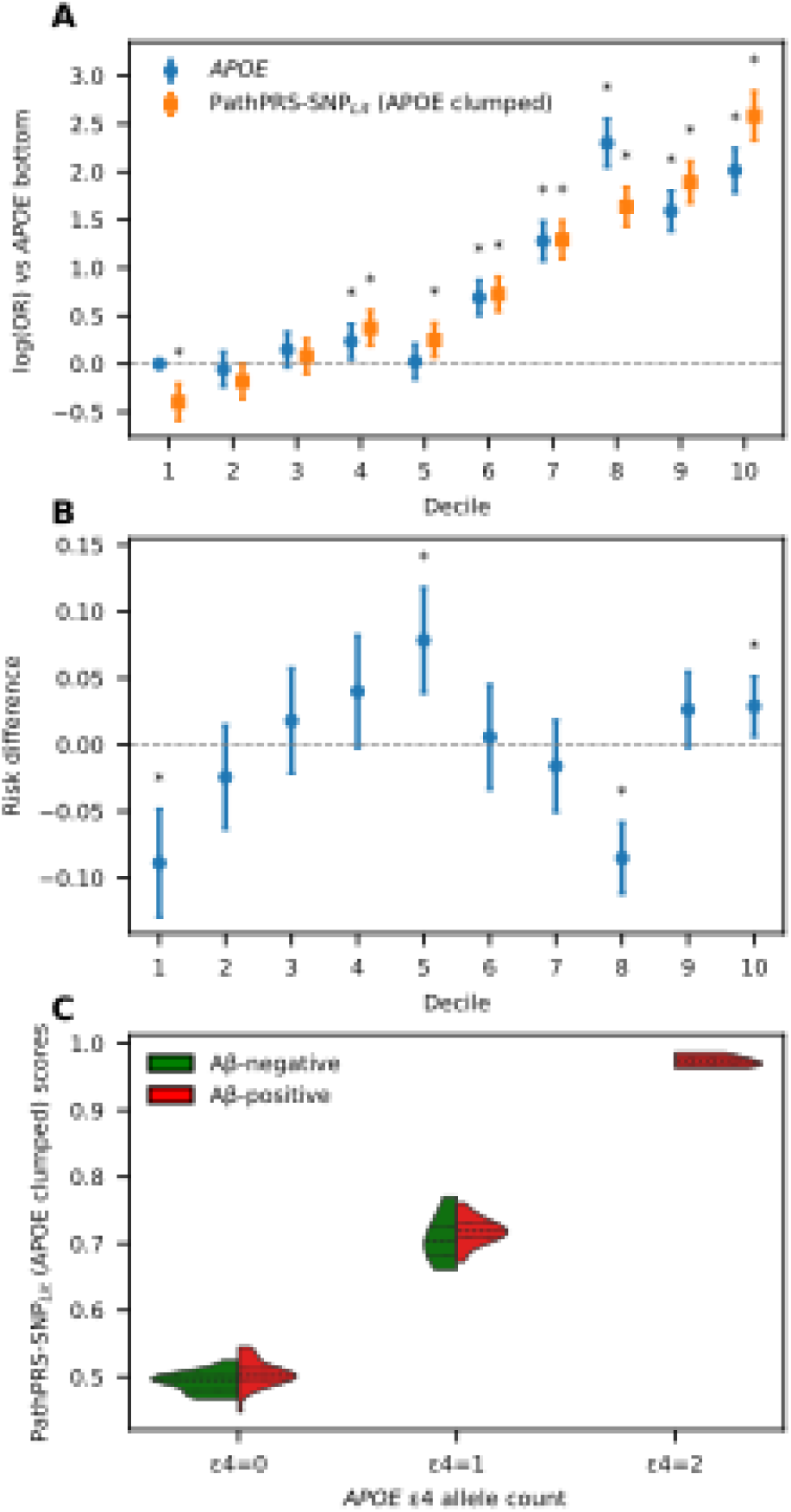
Meta analysis decile-and genotype-based comparison between the *APOE* GRS and the PathPRS-SNP_Lit_ model. (A) Log (ORs) for the *APOE* GRS, referenced to the first *APOE* decile. (B) RD between the two models in each decile. Positive values indicate higher prevalence of predicted abnormal A in PathPRS-SNP_Lit_ than by the *APOE* GRS; error bars give 95% CIs from a meta[analysis across outer[folds. Asterisks (*) mark deciles where the RD differs significantly from 0 (*q*<0.05, two[sided Wald z-test, Benjamini-Hochberg correction). In (A) and (B) error bars show 95% CIs; stars denote deciles whose log-OR differs from the reference with FDR[adjusted *q*<0.05. (C) Violin plot of PathPRS-SNP_Lit_ scores from the fold showing the largest AUC difference between *APOE* and PathPRS-SNP_Lit_. Scores are stratified by *APOE* 4 genotype count (0, 1, or 2), with red and green halves representing A positive and A negative individuals, respectively. Black horizontal bars indicate the 25th, 50th (median), and 75th percentiles (quartiles) of the score distributions. Odds ratio (OR); Risk difference (RD).

To evaluate the underlying performance of all the models (SVM_rbf_, SVM_linear_, Random Forest, Linear Regression, Lasso, Elastic Net, and MLP) across the different PRS feature sets, we applied all models to feature sets containing more than three features: PathPRS-ENS_Lit_, PathPRS-ENS_DB_, and C+T. The AUC distributions were derived from the outer cross--validation results of 10 repetitions of 10×10 stratified nested cross-validation on the ADNI dataset (Supplementary Fig S1, Supplementary Table S3). Across all feature sets, Elastic Net and MLP delivered the best performance, with only PathPRS-ENS_Lit_ features yielding significant gains over *APOE* GRS. Because Elastic Net achieved performance comparable to the MLP while using a simpler model structure, we focused on Elastic Net models in subsequent analyses. Following the principle of Occam’s razor, when two models provide similar predictive performance, the simpler model is generally preferred as it is easier to interpret and less prone to overfitting.

To evaluate the dependence of model performance on the *APOE*, we performed a sensitivity analysis excluding *APOE* SNPs (rs429358, rs7412) and SNPs in linkage disequilibrium r^2^ larger than 0.1 with *APOE* SNPs (rs429358, rs7412). Under this setting, the performance of the PathPRS Elastic Net model decreased substantially (median AUC: 0.758 vs. 0.725, *p* = 9.6×10^-6^), whereas PathPRS-SNP maintained relatively stable performance (median AUC: 0.759 vs. 0.763). This result suggests that the Elastic Net aggregation of pathway-level PRS partly relies on *APOE*-related signals distributed across multiple pathways. In contrast, the SNP-level representation in PathPRS-SNP appears more robust to the removal of *APOE*-linked variants. Therefore, genetic variants outside the *APOE* region can provide additional and independent predictive value.

### PathPRS Outperforms *APOE* in Patient Discrimination

Because PathPRS-SNP_Lit_ is a simpler model with only three features as compared with PathPRS-ENS_Lit_, we selected it for the stratification analyses. Since *APOE* is the established genetic baseline for AD risk prediction, we first evaluated its ability to stratify individuals by risk deciles and used it as the primary comparator for PathPRS-SNP_Lit_ with *APOE* clumped. In genetic testing, the most clinically relevant goal is to distinguish the highest-risk and lowest-risk individuals, rather than focusing solely on overall discrimination as measured by AUC. We therefore examined how each model separated risk across deciles. These analyses demonstrate that PathPRS-SNP_Lit_ provides more informative risk stratification than *APOE* alone, particularly in distinguishing between individuals at the extremes of genetic risk.

We conducted a meta-analysis across 100 models from the outer folds of 10 repeats of 10×10 stratified nested cross--validation (Fig3). In the decile-based meta-analysis, both the *APOE* and PathPRS-SNP_Lit_ models showed a monotonic increase in risk across deciles when referenced to the bottom *APOE* decile. The *APOE* model exhibited significant enrichment only from the fourth decile onwards (*p* = 0.011), whereas PathPRS-SNP_Lit_ already distinguished the bottom decile with a significantly lower odds ratio (OR = 0.67, 95% CI: 0.56–0.81, *p* = 4.5 × 10^−5^). Moreover, PathPRS-SNP_Lit_ identified significantly elevated risk beginning at the fourth decile (OR = 1.45, 95% CI: 1.21-1.75, *p* = 6.5 × 10^−5^), with subsequent deciles showing robust significance (*p* < 0.01). At the top decile, the PathPRS-SNP_Lit_ model reached an OR of 13.23 (95% CI: 10.23–17.11), exceeding the corresponding *APOE* estimate of 7.50 (95% CI: 5.95–9.45), (Supplementary Table S4), (Supplementary Table S5). These results indicate that PathPRS-SNP_Lit_ provides stronger risk stratification than *APOE* alone, particularly in both the low-and high-risk tails of the distribution.

In the risk difference (RD) meta-analysis comparing PathPRS-SNP_Lit_ with *APOE* across deciles, the models showed notable divergence at both ends of the distribution (Fig3). PathPRS-SNP_Lit_ classified substantially fewer individuals as positive in the lowest decile (RD = −0.09, *p* = 1.7 × 10^−5^), while capturing markedly more positives in the fifth decile (RD = 0.08, *p* = 9.97 × 10^−5^). Additional differences were observed in the eighth decile, where PathPRS-SNP_Lit_ assigned lower risk compared with *APOE* (RD = −0.09, *p* = 3.4 × 10^−10^), and in the top decile, where it assigned higher risk (RD = 0.03, *p* = 0.012). These results indicate that PathPRS-SNP_Lit_ provides sharper separation of risk, particularly by down-weighting low-risk individuals and amplifying risk in mid-to-high deciles. The complete decile-wise results, reporting RD, log (OR), and OR estimates with 95% confidence intervals and *p* for each decile are presented in Supplementary Table S6.

In the *APOE* [4 genotype-stratified analysis of PathPRS-SNP_Lit_ scores, brain Aβ-positive individuals consistently showed higher scores than brain Aβ-negative individuals within each *APOE* [4 subgroup (Fig3). For *APOE* [4 non-carriers, the difference was statistically significant (median 0.502 vs. 0.495, *p* = 0.007), indicating that PathPRS-SNP_Lit_ captures risk variation beyond *APOE* [4 status. A similar trend was observed among *APOE* [4 heterozygotes (median 0.718 vs. 0.705, *p* = 0.42), although it did not reach statistical significance. For *APOE* [4 homozygotes, all available individuals were brain Aβ-positive, precluding formal comparison. These results suggest that PathPRS-SNP_Lit_ provides additional discriminatory value within each *APOE* [4 genotype group, particularly among *APOE* [4 non-carriers.

### PathPRS Demonstrates Good Generalization Across an Independent Dataset

The gold standard for assessing model performance is external validation, as an independent cohort provides a clearer indication of whether PathPRS-SNP_Lit_ with *APOE* clumped could be clinically applicable. To ensure consistency in brain Aβ measurements between ADNI and AIBL, we used centiloid values to define brain Aβ positivity. Here, we trained each PRS on the entire ADNI dataset and evaluated its performance on AIBL. PathPRS-SNP_Lit_ continued to outperform baseline models and was particularly effective at identifying individuals with normal brain Aβ levels.

In the external validation on the AIBL dataset (Figure 4), models trained on ADNI demonstrated comparable generalization performance across different PRS inputs. PathPRS-SNP_Lit_ with *APOE* clumped (AUC = 0.694, 95% CI: 0.626–0.762) and PathPRS-SNP_DB_ (AUC = 0.693, 95% CI: 0.626–0.760) both achieved numerically higher discrimination than *APOE* alone (AUC = 0.664, 95% CI: 0.607–0.720), although the differences were not statistically significant (DeLong *p* > 0.15). At fixed false positive rates (FPR), McNemar tests indicate that PathPRS-SNP_Lit_ provided more balanced error control, reducing false positives at low FPR (0.10) and high FPR (0.90), while maintaining comparable sensitivity. These results suggest that pathway--informed PRS generalizes at least as well as *APOE*, with potential advantages in specific operating regions of the Receiver Operating Characteristic (ROC) curve.

**Fig 4:**
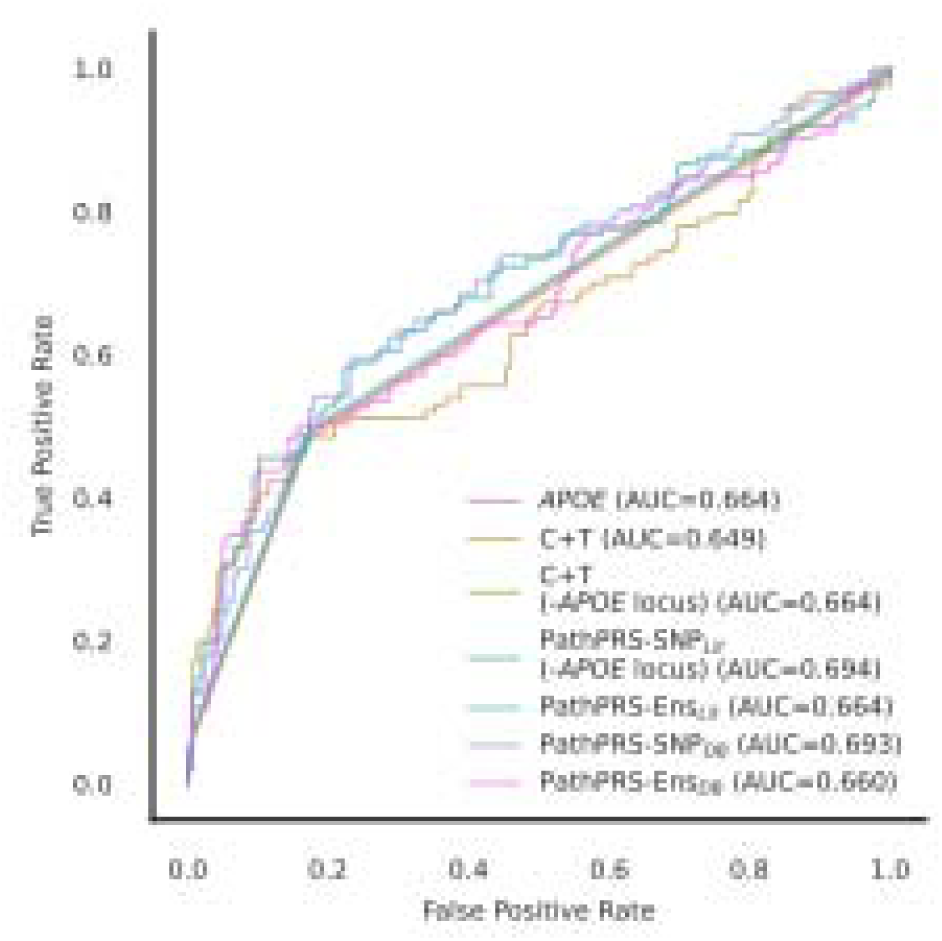
Generalization performance was evaluated on the AIBL dataset. Models were trained on ADNI using different PRS inputs and tested on AIBL.

### Key Pathways Contributing to the Predictive Performance of PathPRS

Since PathPRS-SNP_Lit_ represents the SNP union and does not include pathway-level features, it does not allow for pathway feature importance analysis. As PathPRS-SNP_Lit_ and PathPRS-ENS_Lit_ exhibited comparable predictive performance, we focused on the latter for interpretability. Therefore, we examined feature importances using the PathPRS-ENS_Lit_ model, which can better elucidate the underlying biological mechanisms.

In the feature importance analysis of the PathPRS-ENS_Lit_ model (Fig 5), *APOE* [4 displayed the largest positive coefficient, consistent with its well-established role in driving Aβ deposition. Beyond *APOE*, several pathways were also identified as significant contributors, including plasma lipoprotein particle assembly, the immune system, protein-lipid complex assembly, protein-lipid complex subunit organization, and endocytosis pathways (all *p* < 1.0 × 10^−45^). The involvement of plasma lipoprotein particle assembly pathway underscores the central role of lipid metabolism and cholesterol transport, processes directly linked to *APOE* function.^36^ The immune system pathway points to the importance of inflammatory and microglial responses in modulating Aβ accumulation.^37^ Both protein-lipid complex assembly and subunit organization relate to membrane stability and lipid transport, which may influence Aβ aggregation and clearance.^38^ Finally, the identification of endocytosis highlights the relevance of vesicular trafficking and receptor-mediated uptake in Aβ processing.^39,40^ Together, these results indicate that PathPRS-ENS_Lit_ captures polygenic risk not only through *APOE* but also via biologically coherent pathways implicated in lipid regulation, immunity, and cellular trafficking.

**Fig 5:**
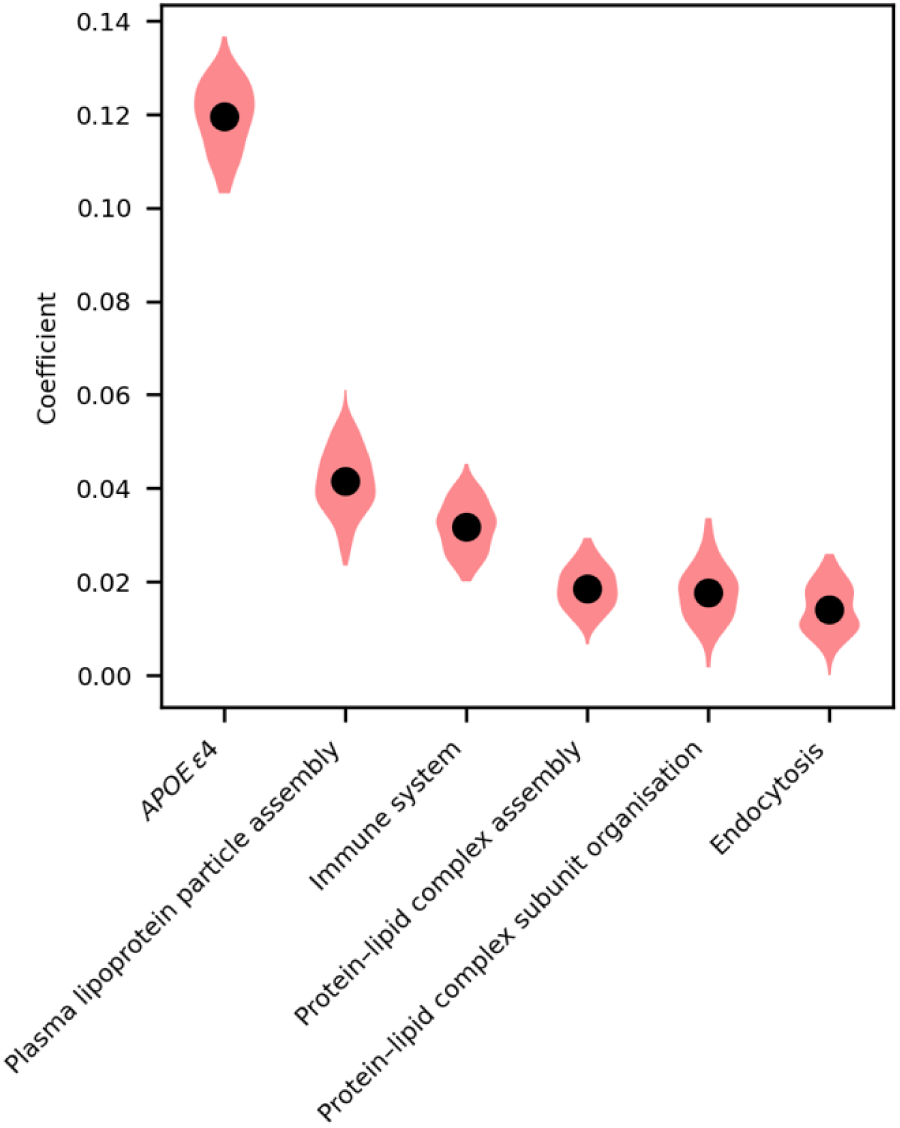
Violin plot of Elastic Net coefficients for the six most influential features in PathPRS-Ens_Lit_. The single violin displays the fold-wise coefficient distribution: the black dot indicates the median and the whiskers show the 95% confidence interval. Coefficients were estimated over all outer folds of the repeated stratified cross-validation and compared with zero using a two-sided one-sample t-test. Resulting p-values were adjusted for multiple testing with the Benjamini-Hochberg FDR procedure; an asterisk marks features that remain significant at FDR < 0.05.

The genes included in PathPRS-ENS_Lit_ and the C+T PRS are listed together with their pathways (Supplementary Table S7), showing that the PathPRS-ENS_Lit_ genes are more tightly linked to AD-related biological functions. This convergence in a small subset of genes suggests that while C+T captures general statistical associations, PathPRS-ENS_Lit_ systematically prioritizes biologically meaningful mechanisms.

Looking at genes mapping to ≥3 pathways in the PathPRS-ENS_Lit_ pathway list, we identified 48 genes (Supplementary Table S7). As expected, this set included well-established AD-risk genes such as *APOE*, *CLU*, *BIN1*, *ABCA7*, *FERMT2*, and additional lipid-related genes including *APOA2*, *APOA4*, *APOB*, *APOC1*, and *APOC3*, along with several others. Of these, 34 were not present in the overlapping C+T gene list, highlighting their potential role as functional hub genes whose associated variants contribute to pathway-level biology. Their contribution to overall risk arises from the combined effects of these variants within each pathway (Fig 5). By restricting analysis to biologically relevant pathways rather than genome-wide SNPs, PathPRS-ENS_Lit_ enhances signal-to-noise, improving both interpretability and predictive performance.

## DISCUSSION

In this study, we set out to systematically evaluate pathway-informed PRS strategies, with Aβ prediction serving as a representative application. Notably, the PathPRS approach, which leverages a carefully selected set of literature-derived pathways, yielded a significantly higher AUC than both the *APOE* GRS and standard C+T PRS in the ADNI cohort. Numerous prior studies have demonstrated that conventional PRS methods such as C+T often yield limited predictive gains over *APOE* GRS when applied to AD biomarkers or clinical diagnosis.^8,25^ Our results confirm this limitation and demonstrate that literature--derived pathways offer greater improvement. Moreover, we observed improved risk stratification at both extremes of the risk spectrum, highlighting the potential clinical value of incorporating pathway-level biological insights into AD risk prediction.

Existing pathway-informed PRS studies use heterogeneous pathway definitions, leading to inconsistent conclusions about their utility for Aβ prediction.^5,8^ In this work, we unified these pathways using machine learning and demonstrated its utility. Notably, several pathways previously considered uninformative when evaluated in isolation,^5^ such as plasma lipoprotein particle assembly and protein-lipid complex assembly, emerged as top contributors in the PathPRS-Ens_Lit_ model. This reveals a key insight: some pathways only demonstrate predictive value when considered alongside others. Complex disease risk is better captured through the combined effects of multiple pathways. These findings highlight the importance of modelling pathway PRS jointly.

A key finding is the marked performance difference between literature-curated pathways (PathPRS_Lit_) and database-derived pathways from FUMA (PathPRS_DB_). The literature-derived pathways, although relatively few in number (32 in total), appear to provide a higher signal-to-noise ratio. In contrast, the much larger set of FUMA-derived pathways (201 pathways) did not confer additional predictive value beyond *APOE* alone. Similar challenges have been reported in previous studies that relied on enrichment analyses via MAGMA or FUMA, where overly broad or noisy gene sets diluted predictive signals.^27^ This discrepancy may stem from the fact that our literature-based pathways, by virtue of being curated more vigorously, are highly informative.

A related contrast arises when comparing PathPRS-SNP_Lit_ to the standard C+T model. Unlike the C+T approach, which selects SNPs solely based on their genome-wide statistical significance under a defined *p* threshold (e.g., *p* < 1×10^−4^), PathPRS-SNP_Lit_ includes SNPs based on their presence in a curated set of literature-derived pathways relevant to AD biology. Despite the fundamentally different SNP selection strategies, the two models share 30 overlapping genes, including established AD risk loci such as *APOE*, *CLU*, *ABCA7*, and *EPHA1* (Supplementary Fig S2), (Supplementary Table S7). This overlapping gene set is enriched in pathways related to lipid metabolism, immune response, and amyloid processing, mechanisms central to AD pathogenesis.^3^

Our evaluations use Aβ positivity as the case study, but the modelling strategies and insights are broadly transferable to other diseases with polygenic architecture. Pathway-informed SNP selection can help isolate genetic variants that are more biologically plausible, thereby improving interpretability and potentially enhancing prediction. Similar approaches have shown promise in cancer^41^ and autoimmune diseases,^13^ where biologically informed PRS models improved interpretability and predictive accuracy. Neurodegenerative diseases like Parkinson’s disease or Amyotrophic Lateral Sclerosis, where discrete pathways are well characterized,^42,43^ could realize even greater benefits.

Beyond the selection of pathways themselves, the broader framework of integrating biological knowledge into PRS is flexible. One could, for instance, incorporate additional functional layers, such as regulatory annotation (e.g., eQTLs) or tissue-specific expression, to further prioritize SNPs. Similarly, more advanced modeling schemes, ranging from graph--based neural networks to nonlinear kernel methods, could be used to capture complex relationships within and across pathways. The present study demonstrates that a relatively simple approach (i.e., aggregating pathway-specific PRSs with a machine learning model) can already outperform standard genome-wide PRSs in a challenging clinical endpoint. Future research could investigate how to unify these integrative strategies into a more comprehensive “network-aware” genetic model.

Several limitations should be noted. First, our focus on individuals of primarily European ancestry restricts the immediate generalizability of these findings to more diverse populations, where LD and allele frequencies differ substantially. Future work can explore pathway-informed PRS in diverse cohorts. Second, the summary statistics used to construct PRS were derived from AD case-control GWAS rather than from GWAS directly targeting Aβ burden. Prior work has suggested that trait-specific summary statistics, such as those from Aβ quantitative GWAS, may yield stronger predictive performance even in the presence of *APOE* effects.^44^ Future studies could therefore investigate whether using trait-specific summary statistics further enhances prediction accuracy.

Looking ahead, a promising line of investigation lies in integrating additional biomarker information (e.g., blood-based Aβ or tau measures) with pathway-level genetic scores for a multimodal risk assessment. Emerging efforts integrating imaging, blood-based biomarkers, and genetic data have shown promise for multimodal AD risk stratification.^45,46^ Integrating these with biologically prioritized PRS may further enhance early detection. Machine learning methods could also be extended to incorporate epistatic interactions within pathways. Finally, while this study focused on early pathology via Aβ positivity, similar strategies could be adapted to other hallmark AD processes, such as tau pathology or neurodegeneration, offering broader coverage of AD’s multifaceted progression.

## Acknowledgements

Data collection and sharing for this project were funded by the Alzheimer’s Disease Neuroimaging Initiative (ADNI; National Institutes of Health grant U01 AG024904) and DOD ADNI (Department of Defense award W81XWH-12-2-0012). ADNI is supported by the National Institute on Aging, the National Institute of Biomedical Imaging and Bioengineering, the Canadian Institutes of Health Research, and the Foundation for the National Institutes of Health, as well as multiple industry partners. The grantee organization is the Northern California Institute for Research and Education, with study coordination by the Alzheimer’s Therapeutic Research Institute and data dissemination by the Laboratory for Neuro Imaging at the University of Southern California.

Data used in the preparation of this article were also obtained from the Australian Imaging, Biomarkers and Lifestyle (AIBL) study. The authors thank the AIBL investigators and participants, and acknowledge the contributions of CSIRO, Edith Cowan University, the University of Melbourne, and the Florey Institute of Neuroscience and Mental Health. AIBL has been supported by CSIRO, the Science and Industry Endowment Fund, the National Health and Medical Research Council (NHMRC), the Dementia Collaborative Research Centres, and industry partners. Genetic data used in this study were supported by NHMRC grants GNT1161706 and GNT2001320 awarded to S.M.L.

## Funding Declaration

This research was supported by the Australian Research Council Training Centre in Cognitive Computing for Medical Technologies (IC170100030) and funded by the Australian Government. The Florey Institute of Neuroscience and Mental Health acknowledges the support of the Victorian Government through the Operational Infrastructure Support Program.

Large language models have been used for grammatical polishing of this paper.

## Author Contributions

XZ contributed to the conception and design of the study, performed data processing and statistical analyses, generated all figures and visualizations, and drafted and revised the manuscript.

BG conceived and designed the study, supervised the analytical framework and methodological development, and contributed to critical revision of the manuscript.

SML contributed to data acquisition, and assisted with revision of the manuscript.

CLM provided domain expertise in AD and contributed to interpretation of findings and manuscript revision. TB contributed to study design, methodological feedback, and manuscript editing.

NGF supervised the project, contributed to study conception, interpretation of results, and revision of the manuscript. Conflicts of Interest

Nothing to report.

## Data Availability

The data that support the findings of this study from ADNI are available upon request in https://adni.loni.usc.edu/data-samples/adni-data/#AccessData and to AIBL data through https://aibl.org.au/collaboration/#data-access. The corresponding author can provide additional guidance on request.

## Biographical Note

Xiyuan (Emma) Zhang is a PhD candidate at The University of Melbourne and the Florey Institute, focusing on machine learning, genomics, and Alzheimer’s disease biomarker prediction.

