## Supplementary Material 1 for "Comparing Pathway-Informed Polygenic Risk Score Strategies: A multi-cohort evaluation of Amyloid-β"

**Supplementary figures**

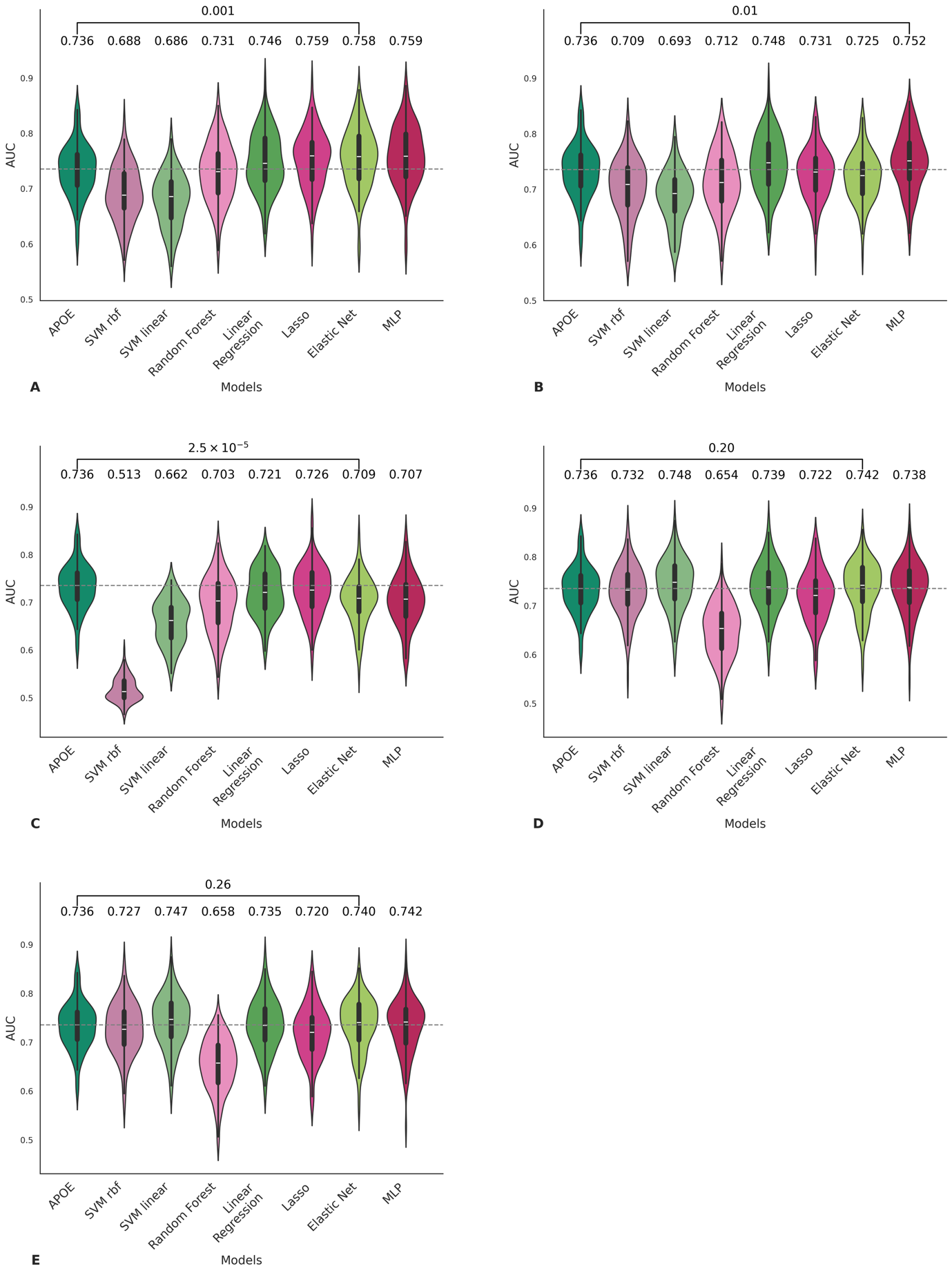

***Supplementary Fig 1: AUC distributions from 10 repeats of*** $\boldsymbol{10}\boldsymbol{\times}\boldsymbol{10}$ ***nested stratified cross‑validation on models trained with different PRS inputs in the ADNI dataset.*** *(A) PathPRS-Ens_Lit_ evaluated with different models; (B) PathPRS-Ens_Lit_ evaluated with different models after excluding the APOE locus; (C) PathPRS-Ens_DB_ evaluated with different models; (D) C+T PRS evaluated with different models; and (E) C+T PRS evaluated with different models after excluding the APOE locus. The distributions were compared using the Mann-Whitney U test. The grey dashed lines indicate the median AUC of the APOE GRS across repetitions.*

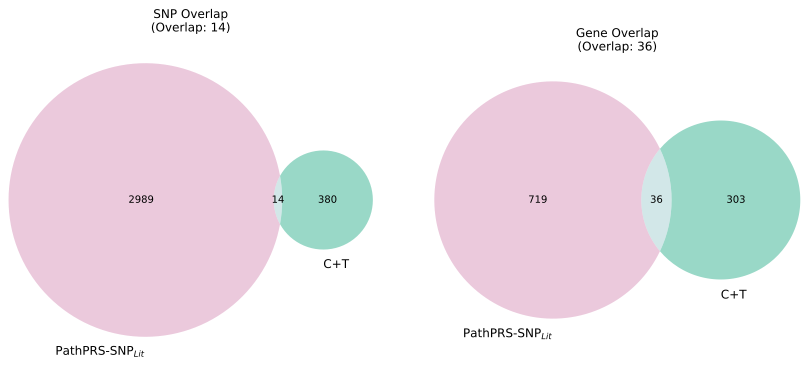

***Supplementary Fig 2: Venn diagram showing the overlap of genes included in the PathPRS-SNP_Lit_ and C+T models, based on SNPs selected using a p-value threshold of*** $\boldsymbol{1\times}\boldsymbol{10}^{\boldsymbol{-4}}$***.***
